# Clinical and haematological profile of dengue among adult patients at a tertiary care hospital in Pokhara

**DOI:** 10.1101/2022.04.19.22274055

**Authors:** Durga Dhungana, Bidhya Banstola, Mahesh Banjara

## Abstract

**Introduction:** Dengue is an important infectious disease. This disease is prevalent in the terai belts of Nepal mainly. But in the last few years, the cases are in increasing trend in the hilly areas of Nepal also. Hence this study was done in an aim to study the clinical and haematological profile of the dengue cases.

**Methods:** This was a retrospective cross-sectional quantitative study done at a tertiary teaching hospital of Pokhara, Nepal after obtaining ethical approval from the institutional ethical committee. The data of serologically confirmed dengue cases, during the period of August 2019 to December 2019, of age above 15 years, were collected and analysed using SPSS 20. Descriptive analysis in terms of mean, median, percentage as well as t-test for nominal and chi-square tests were used to compare different parameters. P-value≤ 0.05 was considered statistically significant.

**Results:** Out of 922 patients, approximately one-half (50.5%) cases were seen during the month of September. Most (82.8%) were the inhabitants of Kaski district. Median age of presentation was 29 years with slightly more cases of males (52.4%). Three hundred and forty seven patients were admitted. Fever (96.5%) and headache (40.6%) were the most common symptoms on presentation of admitted cases. Leukopenia (55.3%) was more common than thrombocytopenia (47.6%) in the admitted cases. On comparison between admitted patients with warning signs and those without signs, no significant variation was seen in terms of age, total leukocyte count and total platelet count.

**Conclusion:** Dengue is common in young population. Fever, headache and gastrointestinal symptoms are common among dengue patients. Leukopenia and thrombocytopenia are common laboratory features of dengue.

## Introduction

Four dengue virus (DENV) types are recognized and correspond to four distinct virus species based on antigenic and genetic characteristics. In tropical areas, transmission is maintained throughout the year and intensifies at the start of the rainy season, when infected vector mosquitoes are more abundant as higher humidity lengthens their life span and increased temperatures shorten the extrinsic incubation period. Dengue occurs in all age groups. DENV transmission and increasing global burden of disease is being fuelled by population growth, urbanization, increasingly favourable ecologic and environmental habitats for Aedes mosquitoes compounded by poor vector control, and the ease of international travel. Today, Asia and the Western Pacific bear nearly 75% of the global dengue disease burden(1).

Clinical manifestations of dengue can be variable ranging from asymptomatic to undifferentiated fever, dengue fever without warning signs, dengue fever with warning signs and severe dengue (previously as dengue haemorrhagic fever and dengue shock syndrome). Modes of transmission are variable but most common mode is through infected male aedes mosquito bite.(2) The central clinical features of severe dengue are haemorrhagic phenomena and hypovolemic shock caused by increased vascular permeability and plasma leakage. Studies have shown that NS1 protein is present early in dengue virus infection and offers a new technique to diagnose a dengue virus infection based on NS1 antigen capture ELISA. Rapid immunochromatographic tests to detect dengue IgM and IgG have demonstrated high sensitivity and specificity. In the absence of a vaccine, dengue prevention currently relies on public health and community-based control programs(1).

Dengue is an emerging infectious disease in Nepal. The first reported case of Dengue was in 2004 A.D from Chitwan. From 2006 to 2019 A.D, the total number of cases has been 8391(2). In regards to Gandaki Province, the cases were nil till last few years with one case reported each in year 2073 B.S and in 2074 B.S. A total of 2679 cases were reported in 2019.

Majority cases i.e. 2575 cases were reported from Kaski district alone(3). Hence there has been a substantial increase in number of cases during the last few years. Hence dengue is not limited to the terai belts only but spreading to the valleys and hilly areas too.

Research on dengue has been limited in Nepal and only few papers are available. Limited research mainly focused on terai belts and mainly on serological and demographic characteristics (4-7). This study focuses on all aspects including the laboratory parameters which will help to estimate the morbidity to a deeper extent which ultimately burdens the economical constraint of the individual, society and the country.

## Methods and Materials

This was a quantitative cross-sectional descriptive study done among dengue patients, diagnosed serologically with positive NS1 antigen and/or IgM antibody tests, which were admitted in the medicine ward or had visited Out-Patient department (OPD) of medicine department of Gandaki Medical College Teaching Hospital and Research Center during the period of August 2019 to December 2019. Gandaki Medical College is a 550 bedded tertiary level hospital of Pokhara. It is one of the referral sites for the western region also. The data was collected between August 2020 to October 2020 after obtaining approval from institutional review committee of Gandaki Medical College (Ref no. **063/2077/2078**). Total enumerative sampling method was used. Patient details were obtained from the medical records department for the admitted cases and the laboratory for the out-patient cases. Data was collected with respect to the demographic profile of the patients, laboratory diagnosis methods with the haematological and biochemical investigations done. Proforma included data on age, sex, address, presenting symptoms, signs, area of admission, duration of hospital stay(for admitted cases), outcome(death or discharged) and laboratory tests. Data of OPD cases were complete in regards to age, sex and gender but many had missing laboratory data, hence were not included in analysis of haematological profile. Among admitted cases, all of them had undergone complete blood count. However other laboratory and radiological tests were done on needed basis as seen in data from the medical records section. Laboratory tests included aspartate transaminase (AST), alanine transaminase (ALT) and renal function tests (RFT). Radiological tests included X-ray and ultrasonography. Thrombocytopenia was defined as platelet count less than 150000/mm3 of blood. Leukopenia was defined as leukocyte count less than 4000/mm3 of blood. Cases of dengue were classified as per WHO classification of dengue(8).

Data was entered into Microsoft excel and checked for repetition. Data regarding outpatient cases were incomplete in regards to haematological and biochemical profile, hence were included in the demographic analysis only. Data of admitted cases were only analysed in regards to laboratory data. Total of 922 cases were obtained after removing repeated cases matching the address, gender and age. Out of which, 347 cases were admitted. Data was then imported into Statistical Package for the Social Sciences (SPSS) 20 and then analysis was done. Descriptive analysis was done for age, gender and address. Further analysis of the admitted cases was done regarding the co-morbidities, clinical features and haematological parameters. T-test for nominal and chi-square tests were used to compare different parameters among admitted cases with regards to differences between dengue with warning signs and dengue without warning signs. P-value≤ 0.05 was considered statistically significant.

## Results

Majority of the cases were seen in the month of September. Median age of presentation was 29 years of age with maximum from Kaski district. (Table 1)

**Table 1.**
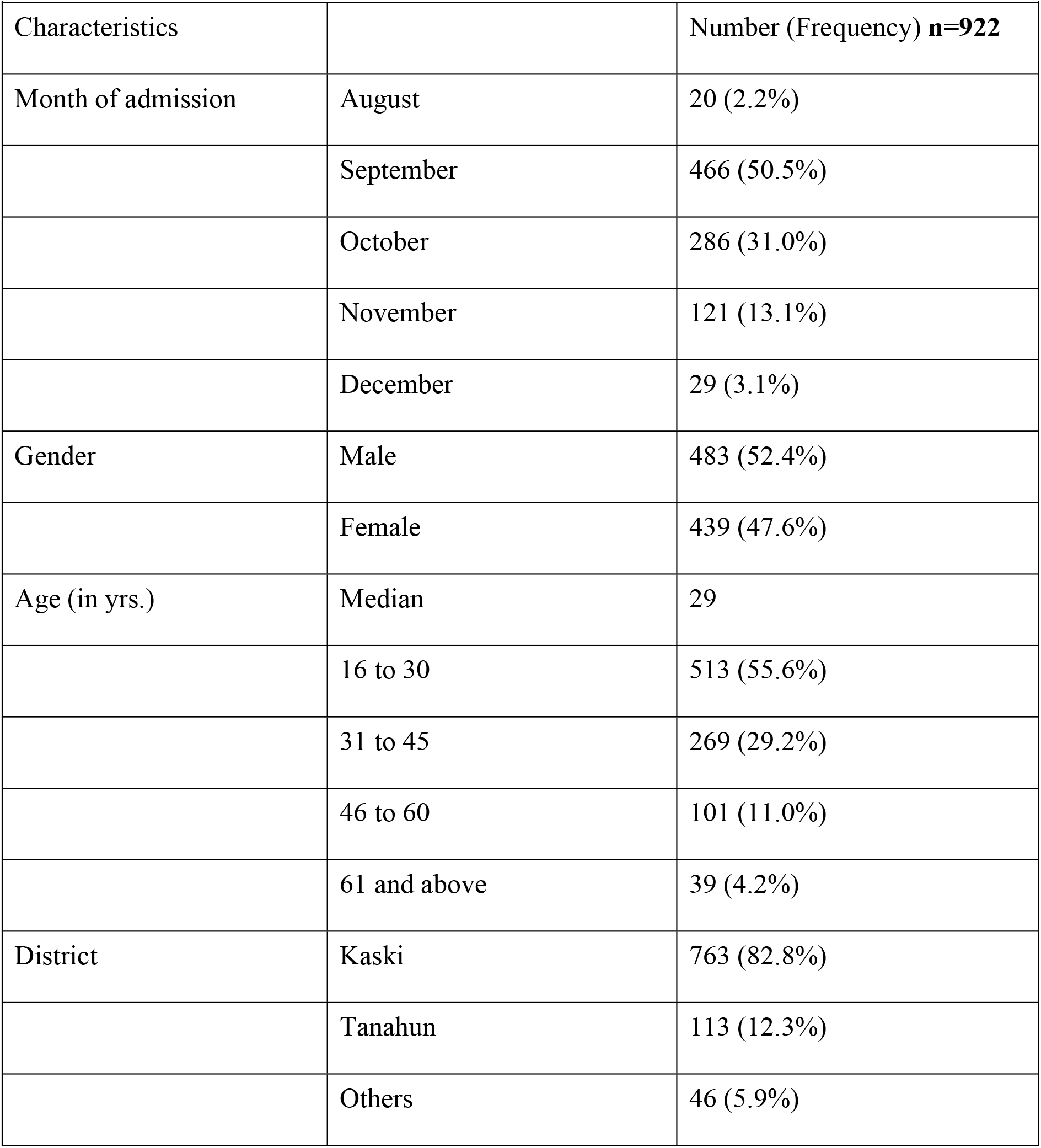
Sociodemographic characteristics

Except three cases, all cases were managed at ward level. Nearly one-ninth of the admitted cases had some known comorbidities. Majority of them were hypertensive. (Table 2)

**Table 2.** Clinical characteristics of admitted cases n=347.

| **Table 2.Clinical characteristics of admitted cases n=347** |  |  |
| --- | --- | --- |
| Characteristics |  | Number (Percentage) |
| Gender | Male | 178 (51.3%) |
|  | Female | 169 (48.7%) |
| Comorbidities | Yes | 38 (11.0%) |
|  | Hypertension | 16 (4.6%) |
|  | Diabetes | 8 (2.3%) |
|  | Pregnancy | 4 (1.1%) |
|  | Others | 13 (3.7%) |
| Lab Diagnosis | NS1 Antigen positive | 337 (97.1%) |
|  | IgM Antibody positive | 5 (1.4%) |
|  | Both NS1 and IgM positive | 5 (1.4%) |
| Admission | ICU | 3 (0.9%) |
|  | Ward | 343 (99.1%) |
| Hospital Stay Duration | ≤ 4 days | 291 (83.9%) |
|  | > 4 days | 56 (16.1%) |

Fever was present in almost all of the admitted cases with gastrointestinal system as most common associated systemic involvement. (Table 3)

**Table 3.** Presenting symptoms among admitted cases n=347.

| **Table 3. Presenting symptoms among admitted cases n=347** |  |
| --- | --- |
| Symptoms | Number (Frequency) |
| Fever | 335 (96.5%) |
| Headache | 141 (40.6%) |
| Gastrointestinal | 104 (30%) |
| Nausea or Vomiting | 80 (23.1%) |
| Diarrhoea | 13 (3.7%) |
| Abdominal pain | 11 (3.2%) |
| Myalgia | 66 (19.0%) |
| Retro orbital pain | 23 (6.6%) |
| Rash | 16 (4.6%) |
| Bleeding | 11 (3.2%) |

Leukopenia and thrombocytopenia were common findings. (Table 4)

**Table 4.** Hematological Parameters of Dengue Patients n=347.

| **Table 4. Hematological Parameters of Dengue Patients n=347** |  |  |  |
| --- | --- | --- | --- |
| Parameter | Value | Minimum | Maximum |
| WBC | 3800 | 1190 | 25930 |
| Leukopenia | 192 (55.3%) |  |  |
| Platelet | 156000 | 14000 | 1349000 |
| Thrombocytopenia | 165 (47.6%) |  |  |

On comparison between patients with warning signs and those without signs, no significant variation was seen in terms of age and other lab parameters. (Table 5)

**Table 5.** Comparison of parameters among those without and with warning signs.

|  |  |  |  |
| --- | --- | --- | --- |
| Parameters | Dengue with warning signs (n=95) | Dengue without warning signs (n=252) | p-value |
| WBC count | 4405 (2565) | 4509 (2463) | 0.73 |
| Age (Mean, S.D) | 31.9 (12.9) | 31.5 (12.8) | 0.81 |
| Platelet count | 160400 (70636) | 178290 (107591) | 0.13 |
| **Categorical** |  |  |  |
| Leukopenia | 56/95 | 136/252 | 0.40 |
| Thrombocytopenia | 48/95 | 117/252 | 0.49 |
| Hospital stay (days) | 3.3 (1.48) | 3.1 (1.68) | 0.32 |

## Discussion

This data on the outbreak of dengue in Pokhara found that about 37.6% of the dengue patients got admitted for treatment. Males were slightly more affected than females (M: F=1.1:1) and similar for the admission. This finding was similar to the study done in India, Taiwan and Malaysia (7, 9-11). Most of the patients were from Pokhara as the hospital is in the heart of Pokhara.

The median age of presentation was 29 years. Older age group represented a minority (<5%) of cases. These findings are similar to the study done in tertiary teaching hospitals in Malaysia and Bangladesh during months of July to September 2019.(9, 12) Among the older age group majority (41%) needed admission.

Most of the admitted cases were managed at the hospital ward level with only three cases needing intensive care and one case expired during the course of treatment. Hypertension (42%) and diabetes mellitus (21%) were the commonest comorbidities present. This was similar to the findings of other studies study(12, 13).

Fever (96.5%) was the most common clinical symptom in this study and was the common finding in other studies too (7, 9, 11-20). The second most common symptom was headache (40.6%) which was similar to the findings of other studies (7, 13). Proportion of patients with headache and myalgia were less in this study as compared to the studies which might be due to entry of the main symptom only during admission and ignoring the less troublesome symptoms by the patient in history taking (9, 12, 17, 19, 21).

Gastrointestinal symptoms in the form of nausea, vomiting, abdominal pain or diarrhoea were seen in nearly one-third of cases. Myalgia was reported by nearly one-fifth (19%) of the cases. Bleeding manifestation was uncommon (3.2%). Sixteen patients had rash on presentation

Leukopenia and thrombocytopenia were seen respectively in 55.3% and 47.6% of total admitted cases. Similar findings were seen in a study conducted in a hospital of eastern India(14). Percentage of patients with findings of leukopenia and thrombocytopenia are variable due to the variation in the cut-off values taken on comparison with other studies. With similar cut-off values also, the difference may be likely due to the timing of blood analysis also. Some may have been in febrile phase whereas others may have been in critical phase (12, 13, 18, 19, 22).

With the newer classification of dengue by WHO, the cases with warning signs were ninety-five (27.3%). This was less as compared to the study done in Bangladesh where 45.5 % admitted cases were with warning signs.(12). This difference is probably due to the parameters taken into account. Radiological findings and detailed clinical examination was not present as well as findings that could have developed later haven’t been taken into account. However a study done in India showed only 11% cases with warning signs (20).This variation is probably due to the study in the latter part was done among both admitted and OPD cases; however this and the former study were conducted among only admitted cases.

Age, total leukocyte count and platelet count couldn’t predict the probability of patients to have warning signs. Hence these parameters were not found predictors of severity or worsening of the cases from simple dengue to severe dengue.

The limitations of the study are that it is a single center study and it is of retrospective nature. The serological type of dengue wasn’t investigated. The differentiation between primary and secondary dengue wasn’t done. Also the haematological profile of the outpatient cases couldn’t be included in the full analysis. However the major power of this study is the large sample size.

## Conclusions

Fever is the most common symptom of dengue in adult patients. With the increasing cases in hilly areas of Nepal, community level programs are needed to be implemented for timely decrease in morbidity and mortality due to dengue. Leukopenia and thrombocytopenia are common findings in dengue patients and should be regarded as suggestive factors.

## Data Availability

All data produced in the present work are contained in the manuscript

## Acknowledgments

None

## References

1. Bennett JE, Dolin R, Blaser MJ. Mandell, Douglas, and Bennett’s Principles and Practice of Infectious Diseases E-Book: Elsevier Health Sciences; 2019.

2. EDCD. National guidelines of prevention, control & management of dengue in Nepal. 2019.

3. Health Directorate GP. Annual Health Report 2076/77. In: Development MoS, editor. 2021.

4. Khetan RP, Stein DA, Chaudhary SK, Rauniyar R, Upadhyay BP, Gupta UP, et al. Profile of the 2016 dengue outbreak in Nepal. BMC research notes. 2018;11(1):1–6.

5. Gupta BP, Haselbeck A, Kim JH, Marks F, Saluja T. The Dengue virus in Nepal: gaps in diagnosis and surveillance. Annals of clinical microbiology and antimicrobials. 2018;17(1):1–5.

6. Gyawali K, Rijal R, Regmi S, Sedhai S, Adhikari S. Study on clinical profile of dengue in a tertiary care hospital of Nepal. Journal of Chitwan Medical College. 2018;8(1):54–8.

7. Saud B, Adhikari S, Maharjan L, Paudel G, Amatya N, Amatya S. An Epidemiological Prospective of Focal Outbreak of Dengue Infection in Kathmandu, Nepal. Journal of Clinical Virology Plus. 2022:100063.

8. Organization WH. Comprehensive guideline for prevention and control of dengue and dengue haemorrhagic fever. 2011.

9. Mallhi TH, Khan AH, Adnan AS, Sarriff A, Khan YH, Jummaat F. Clinico-laboratory spectrum of dengue viral infection and risk factors associated with dengue hemorrhagic fever: a retrospective study. BMC infectious diseases. 2015;15(1):1–12.

10. Nelson V, Thomas TP, Stephen S, Simon S. Clinical Profile of Dengue Fever Outbreak in 2017-A Cross-Sectional Study from South Kerala. 2020.

11. Kuo H-J, Lee K, Liu J-W. Analyses of clinical and laboratory characteristics of dengue adults at their hospital presentations based on the World Health Organization clinical-phase framework: Emphasizing risk of severe dengue in the elderly. Journal of Microbiology, Immunology and Infection. 2018;51(6):740–8.

12. Rafi A, Mousumi AN, Ahmed R, Chowdhury RH, Wadood A, Hossain G. Dengue epidemic in a non-endemic zone of Bangladesh: clinical and laboratory profiles of patients. PLoS Neglected Tropical Diseases. 2020;14(10):e0008567.

13. Nair KR, Oommen S, Pai V. Clinico-Hematological Profile of Dengue Fever during the Monsoon of 2016 in Central Kerala. International Journal of Health Sciences & Research (www ijhsr org). 2018;8(8).

14. Murmu M, Singh LK, Kamble SS, Dash L, Karun Mahesh K, Kandathil JJ. Clinico-laboratory profile of dengue patients in a tertiary hospital of Eastern India. International Journal of Research in Medical Sciences. 2018;6(5):1600.

15. Ayyadevara R. Clinical profile of dengue and its effect of on biochemical parameters: A hospital-based cross-sectional study. MRIMS Journal of Health Sciences. 2021;9(4):151.

16. Naik M, Bhat T, Wani AA, Amin A, Jalaalie U. Clinical and Laboratory Profile of Dengue in Kashmir Valley. The Journal of the Association of Physicians of India. 2022;69(12):11–2.

17. Patil PS, Chandi DH, Damke S, Mahajan S, Ashok R, Basak S. A Retrospective Study of Clinical and Laboratory Profile of Dengue Fever in Tertiary Care Hospital, Wardha, Maharashtra, India. J Pure Appl Microbiol. 2020;14(3):1935–39.

18. Goweda R, Faisal A. A STUDY OF CLINICAL FEATURES AND LABORATORY PROFILE OF DENGUE FEVER IN OUTPATIENT SETTING. Malaysian Journal of Public Health Medicine. 2020;20(2):94–100.

19. Jayadas T, Kumanan T, Arasaratnam V, Gajapathy K, Surendran SN. The clinical profile, hematological parameters and liver transaminases of dengue NS1 Ag positive patients admitted to Jaffna Teaching Hospital, Sri Lanka. BMC research notes. 2019;12(1):1–5.

20. Tewari K, Tewari VV, Mehta R. Clinical and hematological profile of patients with dengue fever at a tertiary care hospital–an observational study. Mediterranean journal of hematology and infectious diseases. 2018;10(1).

21. Karoli R, Fatima J, Siddiqi Z, Kazmi KI, Sultania AR. Clinical profile of dengue infection at a teaching hospital in North India. The Journal of Infection in Developing Countries. 2012;6(07):551–4.

22. Ahmed I, Raza F, Iqbal M, Ashraf M. Dengue virus serotypes and epidemiological features of dengue fever in Faisalabad, Pakistan. Tropical Biomedicine. 2017;34(4):928–35.

